# MultiCOVID: a multi modal Deep Learning approach for COVID-19 diagnosis

**DOI:** 10.1101/2023.01.17.23284647

**Authors:** Max Hardy-Werbin, José Maria Maiques, Marcos Busto, Isabel Cirera, Alfons Aguirre, Nieves Garcia-Gisbert, Flavio Zuccarino, Santiago Carbullanca, Luis Alexander Del Carpio, Didac Ramal, Ángel Gayete, Jordi Martínez-Roldan, Albert Marquez-Colome, Beatriz Bellosillo, Joan Gibert

## Abstract

The rapid spread of the severe acute respiratory syndrome coronavirus 2 (SARS-CoV-2) led to a global overextension of healthcare. Both Chest X-rays (CXR) and blood test have been demonstrated to have predictive value on Coronavirus Disease 2019 (COVID-19) diagnosis on different prevalence scenarios. With the objective of improving and accelerating the diagnosis of COVID-19, a multi modal prediction algorithm (MultiCOVID) based on CXR and blood test was developed, to discriminate between COVID-19, Heart Failure (HF) and Non-Covid Pneumonia (NCP) and healthy (Control) patients. This retrospective single-center study includes CXR and blood test obtained between January 2017 and May 2020. Multi modal prediction models were generated using opensource DL algorithms. Performance of the MultiCOVID algorithm was compared with interpretations from five experienced thoracic radiologists on 300 random test images using the McNemar-Bowker test. A total of 8578 samples from 6123 patients (mean age 66 +/- 18 years of standard deviation, 3523 men) were evaluated across datasets. For the entire test set, the overall accuracy of MultiCOVID was 84%, with a mean AUC of 0.92 (0.89-0.94). For 300 random test images, overall accuracy of MultiCOVID was significantly higher (69.6%) compared with individual radiologists (range, 43.7%- 58.7%) and the consensus of all five radiologists (59.3%, P<.001). Overall, we have developed a multimodal deep learning algorithm, MultiCOVID, that discriminates among COVID-19, heart failure, non-covid pneumonia and healthy patients using both CXR and blood test with a significantly better performance than experienced thoracic radiologists.

## 1. Introduction

The outbreak of Coronavirus Disease 2019 (COVID-19), caused by severe acute respiratory syndrome coronavirus 2 (SARS-CoV-2), stroke the worldwide population with more than 200 million cases and 4.5 million deaths by August 2021. The rapid spread of the pandemic led to a global overexertion of health care and research facilities in order to counteract the growing rate of infection. However, a collapse of the sanitary system was imminent and inevitable worldwide, and new technologies were needed to speed up the diagnostic process.

The reference for COVID-19 diagnosis is the detection of SARS-CoV-2 viral RNA by real-time polymerase chain reaction (RT-PCR). However, the massive requests for sample processing at the beginning of the pandemic, caused serious delays to obtain results.

As lung involvement is one of the main causes of morbidity and mortality in SARS-CoV-2 infection, a quick identification of characteristic findings in chest imaging can support the diagnosis and speed up the identification of COVID-19 positive patients at the emergency units.

Several studies have shown that implementation of Deep Learning (DL) tools to detect chest X-rays (CXR) findings typically associated with SARS-CoV-2 infection, deliver comparable results to those acquired by interpretation of radiologists. However, most of the trained models have a drop in their prediction performance when tested over external datasets (DeGrave et al., 2021). In addition, one of the main hurdles to overcome when training an algorithm to detect Sars-CoV-2 infection in CXR is the similarity of findings with other entities like bacterial pneumonias or heart failure (Cleverley et al., 2020). On the other hand, models based on laboratory results of peripheral blood also give predictive results on diagnosis (Avila et al., 2020) and prognosis (Razavian et al., 2020).

A key fact to highlight is how the incursion of COVID-19 caused a dramatic drop on the emergency room consultations of other pathologies. Later on, after the initial peak, the decline of the COVID-19 prevalence made the non-covid diseases emerge once again at the hospitals. This is relevant due to the challenge of performing an efficient differential diagnosis with selected pathologies during a pandemic. It is well known that the predictive value of a diagnostic test is conditioned by the prevalence of the disease and that of COVID varies widely throughout the different waves of the pandemic (Trevethan, 2017). A multicategory approach that takes into account differential diagnoses that are more stable in their prevalence could reduce this variability.

With the objective of improving and accelerating the diagnosis of COVID-19, we developed a tool to assist physicians in reaching a diagnosis. This tool is a multi-modal prediction algorithm (MultiCOVID) based on CXR and blood test with the ability to discriminate between COVID-19, Heart Failure (HF), Non-Covid Pneumonia (NCP) and healthy (Control) samples.

## 2. Materials and Methods

### 2.1 Dataset

We retrospectively collected CXR images and hemogram values from 8578 samples from 6123 patients and healthy subjects (mean age 66 +/- 18 years of standard deviation, 3523 men) from Parc Salut Mar (PSMAR) Consortium, Barcelona, Spain. Four cohorts were designed: (i) 1171 samples from patients diagnosed with COVID-19 by RT-PCR from March to May 2020; (ii) 1008 samples of patients who suffered an episode of heart failure between 2012 to 2019; (iii) 490 samples of patients diagnosed with non-COVID pneumonia (NCP) from 2018 to 2019; (iv) 5909 samples of standard preoperatory studies of healthy subjects from 2017 to 2019 (**Figure 1**). HR and NCP diagnosis were selected as defined by the International Classification of Diseases, Tenth Revision (ICD-10) code. All the CXR images from groups i-iii were validated by two independent radiologists (MB and JM). Data acquisition and analyses were approved by the local ethics committee of PSMAR Consortium (2020/9199/I).

**Figure 1.**
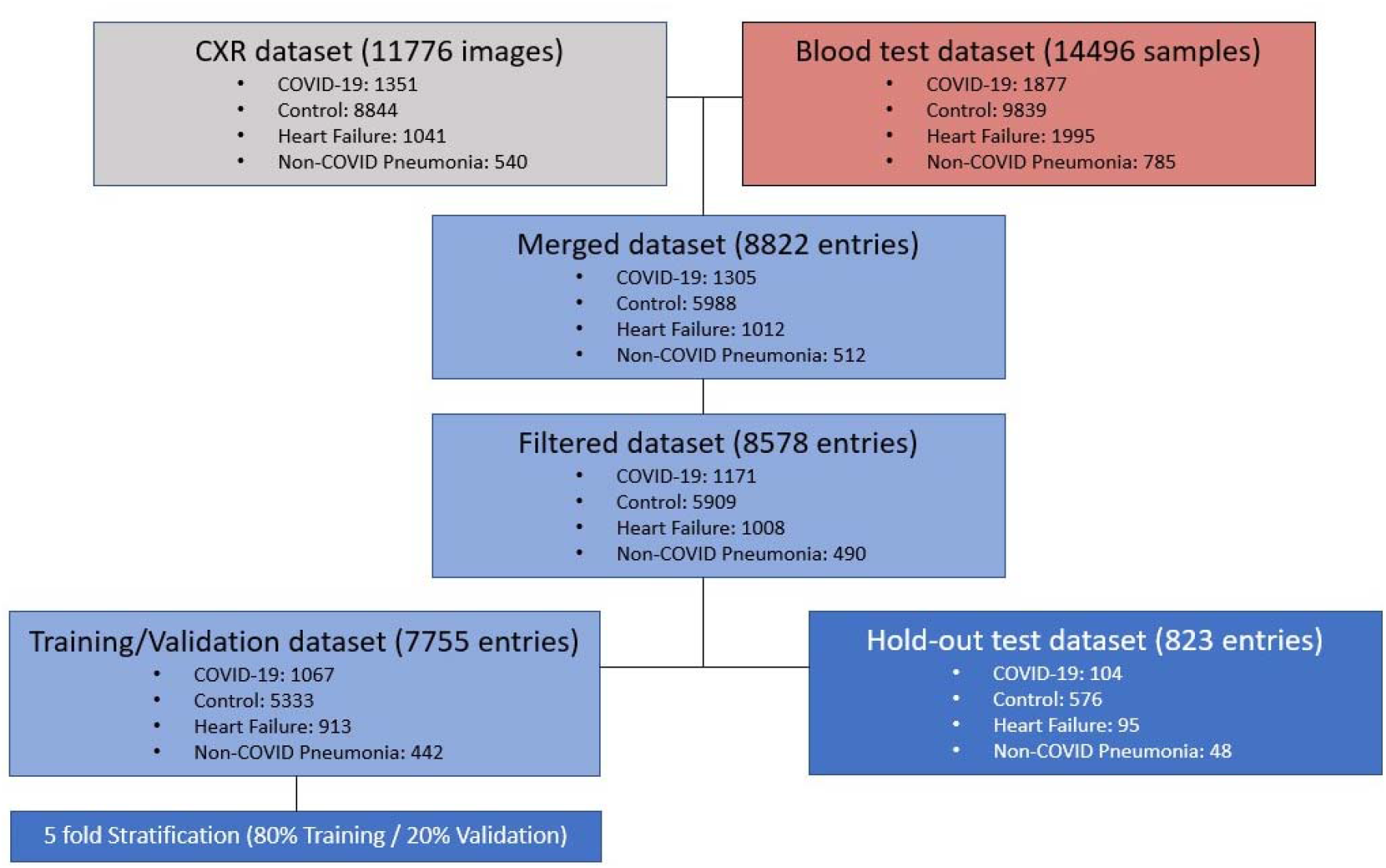
Flowchart for sample selection and patient inclusion in the study and breakdown of training, validation, and hold-out test data sets. Around 25000 entries were obtained using both CXR images and blood test in a time wise manner. The whole dataset totals to 8822 entries of paired CXR and blood test data. Samples with low completeness were discarded for the model building.

### 2.2 Acquisition of blood sample and image data

We included CXR images performed in a period ranging from 1 day before the patient’s diagnosis to 7 days after. The images were filtered to include only frontal projections regardless of the quality and the radiography system used. Blood sample results were collected within a range of 2 days before or 7 days after the CXR acquisition date using PSMAR lab record system, except for control samples whose measurements ranged for two weeks. If two or more blood test results were collected, measurements were averaged.

CXR images and blood test results were combined in the same dataset and split into train (70%), validation (20%) and test (10%) while ensuring that there were no cross-over patients between groups.

### 2.3 Deep learning models

Detailed description of the models, training policy and image preprocessing are provided in Supplementary Material. In brief, segmentation model is based on a U-Net architecture (Ronneberger et al., 2015). The CXR-only classification model consists on a validated Convolutional neural network (CNN) resnet-34 architecture(He et al., 2016). Tabular only-model is an Attention-based network (TabNet) (Arik, S. Ö. and Pfister, 2021). MultiCOVID model is an ensemble prediction of multi-modal deep learning algorithms which merge the CXR-only and the Blood-only and uses both CXR image and blood tests as input values. It uses Gradient Blending in order to prevent overfitting and improve generalization of the joint model (Wang et al., 2019). The whole pipeline development and training was performed using fastai deep learning API (Howard and Gugger, 2020).

Our code base is provided on GitHub at https://github.com/Tato14/MultiCOVID, including weights for each of the individually trained neural network architectures and respective model weights for the weighted ensemble model. Moreover, the anonymized CXR and blood test data is also available upon request.

### 2.4 Comparison with Thoracic Radiologist Interpretations

Hold-out test dataset consisting on 300 samples (ensuring no patient overlap with training or validation sets) was used for expert interpretation. Each sample consisted of a CXR with matched blood results. Expert interpretations were independently provided by five board-certified thoracic radiologists (FZ, SC, LdC, DR, AG) with 2 to 30 years post-residency training experience. Radiologists were able to check both non segmented images and blood test results without any other additional information in a platform created ad-hoc for prediction. They provided a classification for each image in one of the four categories (COVID-19, control, HF and NCP). A consensus interpretation for the radiologist was obtained by the majority vote for each paired CHX-blood test analyzed.

### 2.5 Statistical analysis

A two-tailed t-test P value was reported when clinical and population blood test differences were assessed. McNemar-Bowker test was used to compare model performance against radiologist majority vote using FDR correction. Plotting and statistical analyses were performed using the packages ggplot, ggpubr and rcompanion in R, version 3.6 (R Core Team; R Foundation for Statistical Computing).

## 3. Results

### 3.1 Patient Characteristics

A total of 8578 samples were evaluated across datasets. Patient characteristics and blood test parameters are shown in **Table 1**. A highly significant difference in age was found between the cohort of patients with heart failure (82.8 +/- 10 years) and the other three cohorts (66.0 +/- 16 years for COVID-19 samples, 63.2 +/-18 years for control samples and 67.8 +/-17 years for NCP samples, P < .001 for each comparison) and was not considered as a valid variable for further classification.

**Table 1.** Patient characteristics.

| Characteristic |  | Overall | COVID-19 | Control | HF | NCP |
| --- | --- | --- | --- | --- | --- | --- |
| n |  | 8578 | 1171 | 5909 | 1008 | 490 |
| Age, mean (SD) | [Years] | 66.128<br>(18.353) | 66.013<br>(16.612) | 63.165<br>(18.281) | 82.823<br>(10.731) | 67.786<br>(17.011) |
| Sex, n (%) | H | 5023<br>(58.557) | 677<br>(57.814) | 3598<br>(60.890) | 460<br>(45.635) | 288<br>(58.776) |
|  | M | 3555<br>(41.443) | 494<br>(42.186) | 2311<br>(39.110) | 548<br>(54.365) | 202<br>(41.224) |
| % Basophils | [%] | 0.300<br>[0.200,0.525] | 0.200<br>[0.100,0.300] | 0.400<br>[0.200,0.600] | 0.333<br>[0.200,0.500] | 0.300<br>[0.175,0.500] |
| Total Basophils | [x10<sup>3</sup>/μL] | 0.030<br>[0.020,0.050] | 0.010<br>[0.010,0.020] | 0.040<br>[0.020,0.055] | 0.030<br>[0.020,0.050] | 0.030<br>[0.017,0.060] |
| % Eosinophils | [%] | 0.700<br>[0.100,1.900] | 0.000<br>[0.000,0.300] | 0.950<br>[0.175,2.150] | 0.900<br>[0.200,2.100] | 0.600<br>[0.000,2.200] |
| Total Eosinophils | [x10<sup>3</sup>/μL] | 0.060<br>[0.010,0.160] | 0.000<br>[0.000,0.020] | 0.080<br>[0.020,0.180] | 0.070<br>[0.020,0.160] | 0.060<br>[0.005,0.200] |
| MCH | [pg] | 29.650<br>[28.300,30.900] | 29.400<br>[28.300,30.550] | 29.800<br>[28.433,31.050] | 29.200<br>[27.387,30.700] | 29.600<br>[28.500,30.700] |
| Hematocrit | [%] | 38.000<br>[33.100,42.250] | 40.000<br>[36.600,43.300] | 38.500<br>[33.600,42.700] | 35.200<br>[31.400,39.200] | 32.025<br>[28.512,36.100] |
| Red Blood Cells | [x10<sup>6</sup>/μL] | 4.290<br>[3.710,4.809] | 4.550<br>[4.080,4.935] | 4.360<br>[3.743,4.860] | 3.930<br>[3.494,4.370] | 3.630<br>[3.160,4.070] |
| Hemoglobin | [g/dL] | 12.600<br>[10.800,14.150] | 13.300<br>[12.000,14.433] | 12.900<br>[11.067,14.400] | 11.300<br>[10.000,12.700] | 10.500<br>[9.200,12.100] |
| Leukocytes | [x10<sup>3</sup>/μL] | 9.020<br>[6.771,12.100] | 6.420<br>[5.060,8.860] | 9.450<br>[7.260,12.655] | 8.660<br>[6.894,11.072] | 10.950<br>[7.700,14.781] |
| % Lymphocytes | [%] | 15.350<br>[8.333,25.100] | 14.900<br>[9.475,21.900] | 16.800<br>[8.400,27.600] | 12.700<br>[8.200,18.912] | 10.100<br>[5.763,17.850] |
| Total Lymphocytes | [x10<sup>3</sup>/μL] | 1.310<br>[0.800,2.010] | 0.930<br>[0.688,1.270] | 1.525<br>[0.920,2.240] | 1.070<br>[0.740,1.560] | 1.140<br>[0.605,1.680] |
| MCHC | [g/dL] | 33.233<br>[32.200,34.167] | 33.000<br>[32.100,33.900] | 33.467<br>[32.500,34.400] | 32.300<br>[31.300,33.200] | 32.717<br>[31.600,33.750] |
| % Monocytes | [%] | 7.100<br>[5.300,8.900] | 6.500<br>[4.400,9.000] | 7.100<br>[5.400,8.800] | 8.000<br>[6.237,9.812] | 6.300<br>[4.308,8.600] |
| Total Monocytes | [x10<sup>3</sup>/μL] | 0.635<br>[0.442,0.840] | 0.415<br>[0.290,0.600] | 0.660<br>[0.480,0.870] | 0.690<br>[0.510,0.890] | 0.690<br>[0.440,0.890] |
| % Neutrophils | [%] | 75.200<br>[63.600,84.400] | 77.000<br>[69.100,84.600] | 73.467<br>[60.900,84.100] | 76.800<br>[69.183,82.900] | 81.050<br>[70.925,89.100] |
| Total Neutrophils | [x10<sup>3</sup>/μL] | 6.450<br>[4.400,9.650] | 4.870<br>[3.470,7.095] | 6.673<br>[4.520,10.080] | 6.470<br>[4.950,8.654] | 8.418<br>[5.500,12.360] |
| P-LCR | [%] | 30.700<br>[25.167,36.900] | 30.800<br>[25.300,36.300] | 30.317<br>[24.950,36.500] | 32.858<br>[26.992,38.400] | 32.300<br>[24.000,40.413] |
| PDW | [fL] | 12.600<br>[11.100,14.400] | 12.600<br>[11.075,14.200] | 12.500<br>[11.100,14.300] | 13.000<br>[11.400,14.900] | 12.850<br>[10.800,15.400] |
| Platelets | [x10<sup>3</sup>/μL] | 222.333<br>[173.000,281.000] | 201.000<br>[157.500,267.000] | 227.000<br>[178.000,280.333] | 215.500<br>[168.000,269.000] | 229.500<br>[161.250,373.625] |
| RDW-CV | [%] | 13.900<br>[13.050,15.300] | 13.300<br>[12.600,14.100] | 13.750<br>[13.000,15.100] | 15.325<br>[14.350,17.100] | 14.800<br>[13.813,16.275] |
| RDW-SD | [fL] | 45.100<br>[41.700,49.650] | 43.200<br>[40.600,46.225] | 44.600<br>[41.337,48.850] | 49.833<br>[46.300,54.975] | 48.100<br>[45.100,52.425] |
| MCV | [fL] | 89.000<br>[85.221,92.700] | 88.800<br>[85.700,92.300] | 88.800<br>[85.000,92.400] | 89.950<br>[85.300,94.263] | 90.250<br>[86.806,93.681] |
| MPV | [fL] | 10.700<br>[10.000,11.500] | 10.700<br>[10.100,11.450] | 10.667<br>[10.000,11.400] | 11.000<br>[10.300,11.700] | 10.900<br>[9.950,11.950] |

Abbreviations: COVID-19: coronavirus disease 2019; HF: heart failure; NCP: non-covid pneumonia; MHC: mean corpuscular hemoglobin; MCHC: mean corpuscular hemoglobin concentration; P-LCR: platelet-large cell ratio; PDW: platelet distribution width; RDW: red cell distribution width; MCV: Mean corpuscular volume; MPV: mean platelet volume

### 3.2 Whole CXR models learn spurious characteristics for classification

Previous studies have demonstrated that deep learning (DL)-based algorithms should be rigorously evaluated due to its ability to learn non relevant features in order to increase its prediction accuracy (DeGrave et al., 2021). For this reason, we first developed a segmentation algorithm able to segment lung parenchyma at a 95%-pixel accuracy. Then, after segmentation, we evaluated the accuracy of the algorithms for three complementary datasets: non-segmented images, segmented regions and excluded regions. After few training epochs the three different models achieved nonrandom accuracies between 67% and 74% (**Figure 2A**). However, attention map exploration on the images showed that the different models based their predictions not only inside but also outside of the lung parenchyma (**Figure 2B**).

**Figure 2.**
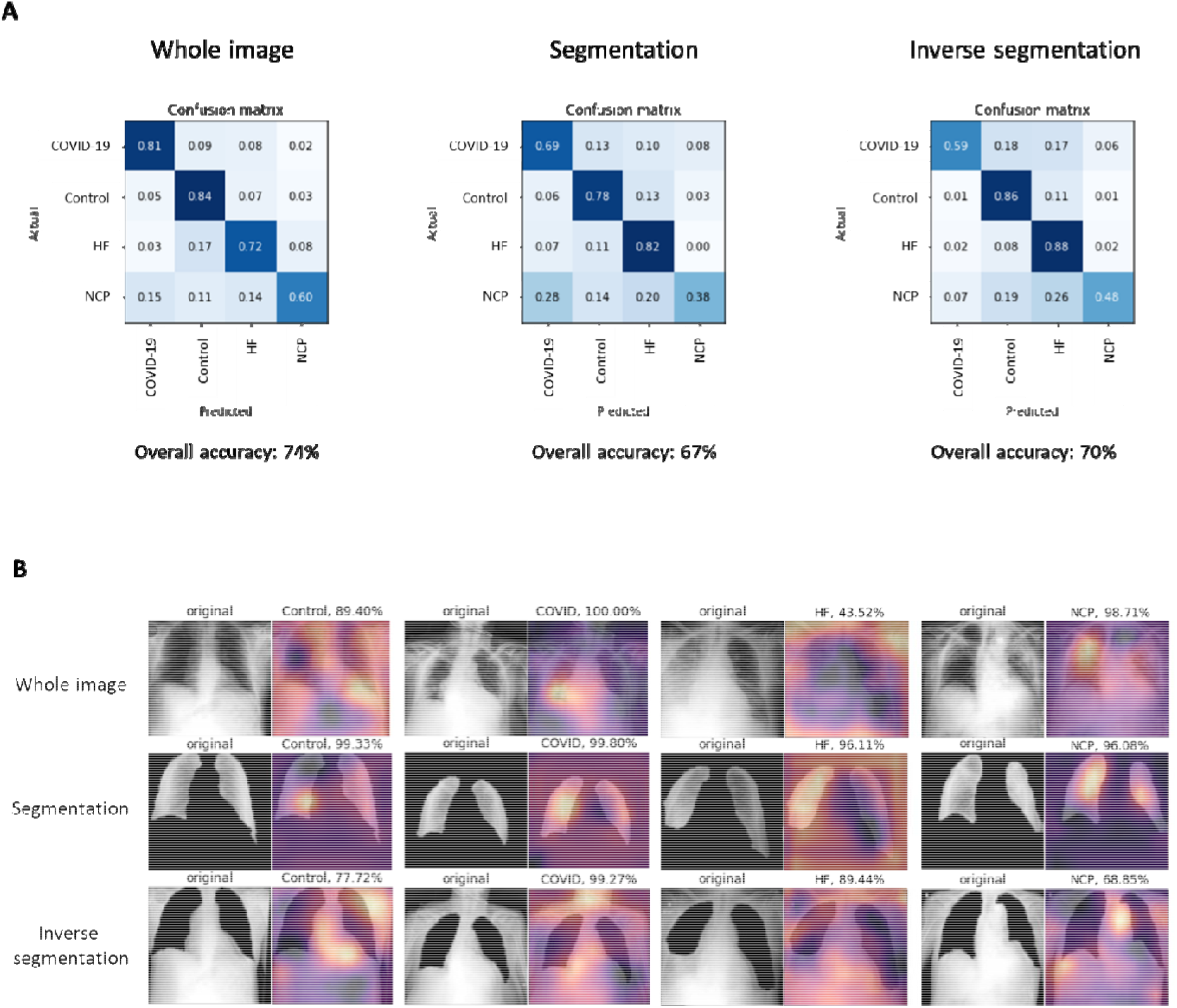
Performance of visual models on whole CXR images. A) Confusion matrix and overall accuracy using whole image, segmented and inverse segmented images, respectively for each category tested. B) Raw image and Grad-CAM heatmap representation of an image for each category and model trained.

These observations showed that our dataset presented fake features that were leading models to correctly classify CXR images. Thus, we decided to first segment all the CXR before training our models for prediction of diagnosis. In order to accomplish this task, we generated a 785-radiology level lung segmentation dataset and trained a U-net model to regenerate the whole CXR dataset keeping only the lung parenchyma.

### 3.3 Performance of single and multimodal models

In order to evaluate the prediction capacity of both segmented CXR and blood sample data, we build different DL models using both sources alone or in combination. Metrics comparison of all the single vision (CXR-only) and tabular (Blood-only) models are detailed in Supplementary Material. As expected, CXR-only models had a more robust prediction of all 4 categories tested compared to Blood-only models (**Figure 3**). This difference is stronger in the classes with less samples (HF, and NCP) where CXR-only models could identify features in the CXR images which are characteristic of these two entities whereas this was not possible with Blood-only models.

**Figure 3.**
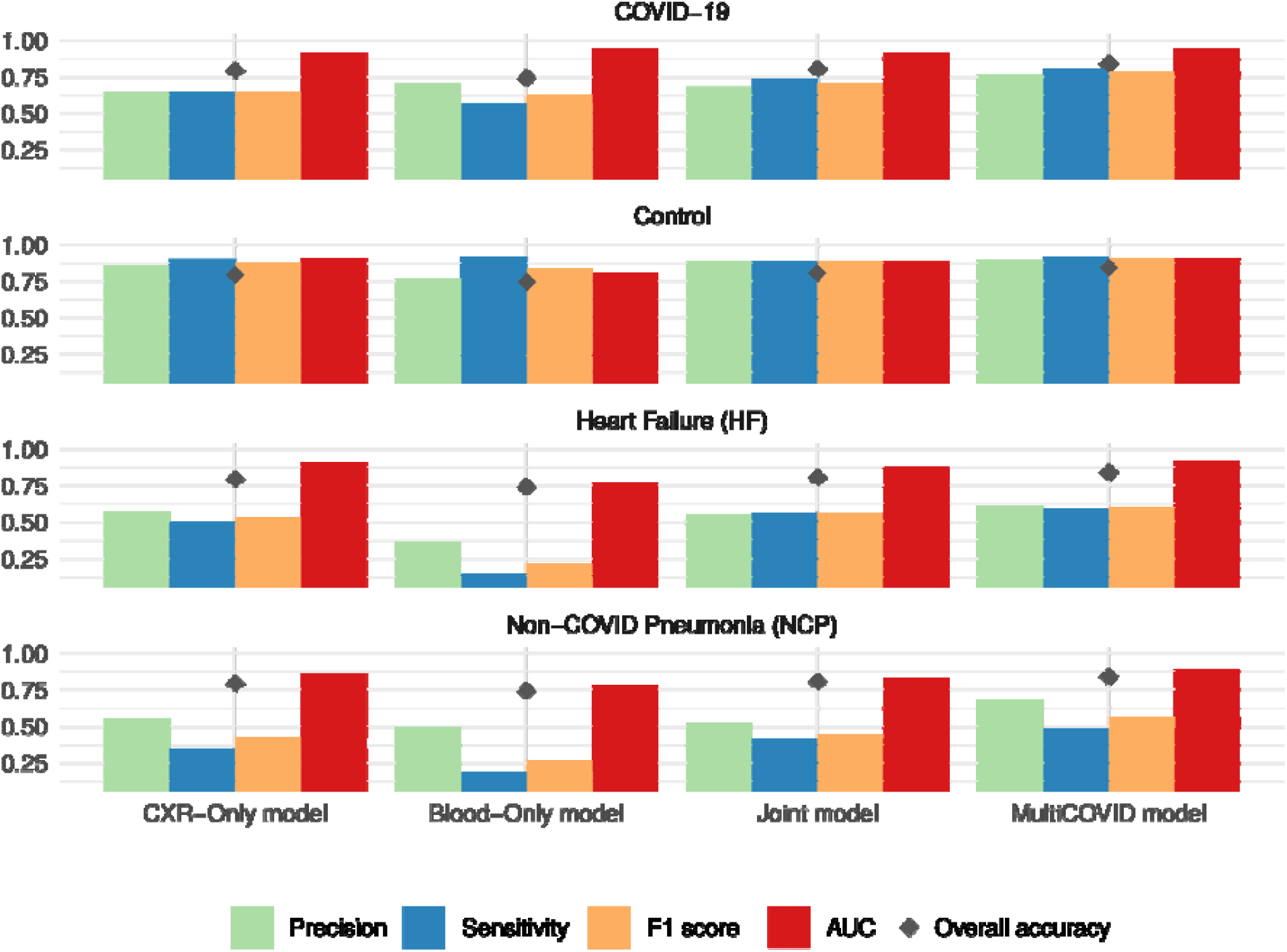
Performance of different models on the entries from hold-out test datasets. Means for precision (green), sensitivity (blue), F1 score (yellow), AUC (red) and accuracy (black diamond) for each model type and category assessed, respectively. CXR-only models use only CXR images for 4 category classification. Blood-only models use blood test a source of information. Joint model uses both CXR and blood test as input for classification and MultiCOVID is the majority vote of 5 different Joint models.

Model interpretability of Blood-only models by analyzing feature importance using Shapley Additive explanations (Lundberg et al., 2020) showed that patient classification was related with two different axes: the immune compartment and the red blood cell (RBC) compartment, respectively (**Figure 4A**). The first axis seems to be strongly associated with COVID-19 classification and shows a specific signature looking at the blood counts (**Figure 4B-top**). However, the second axis seems to subdivide patients between COVID-19/Control and HF/NCP, although COVID-19 blood counts seems to be statistically different from Control samples, too (**Figure 4B-bottom**).

**Figure 4.**
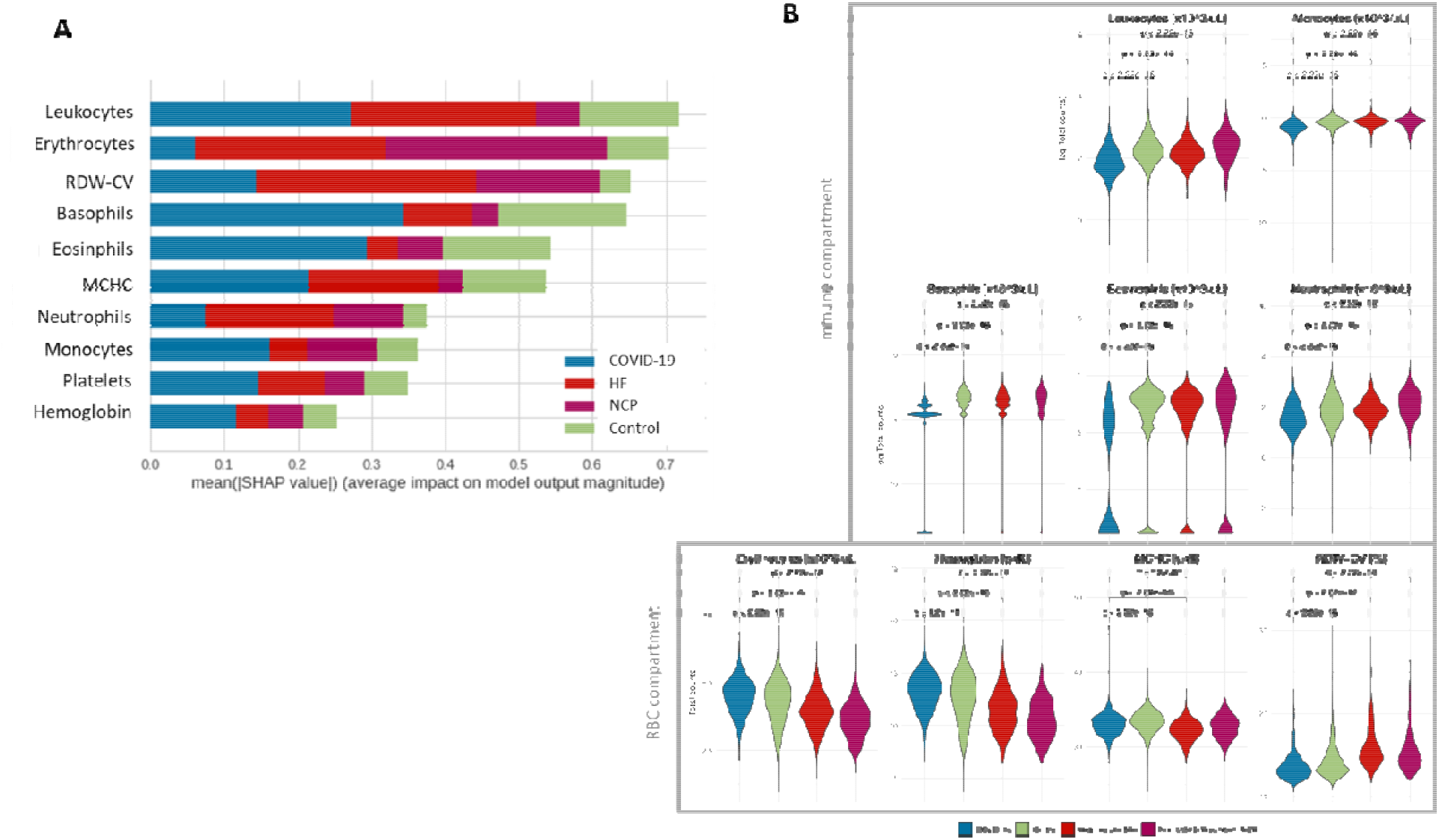
Blood-only model interpretability by SHAP analysis. A) Summary plot showing the mean absolute SHAP value of the ten most important features for the four classes. B) Blood test values of the different features identified by SHAP analysis. RDW-CV: red cell distribution width; MCHC: Mean Corpuscular Hemoglobin Concentration; RBC: red blood cells

The combination of CXR and blood tests using multimodal models that combine inputs from tabular and image data to perform a global prediction, slightly increased the prediction capacity of the single models even when DL tabular models are worse than machine learning (ML - XGBoost) models alone (Supplementary Table). This underpins the concept that adding new sources of information to the data could increase the ability of the models to generate better predictions (Ngiam et al., 2011). Moreover, the joint approach used for building MultiCOVID algorithm resulted on an improved performance in the majority of the metrics analyzed (Figure 3 and Supplementary Table).

### 3.4 Comparison with Expert Thoracic Radiologists

Finally, we compared the performance of MultiCOVID algorithm with the interpretation of expert chest radiologists. This comparison was performed with 300 CXR randomly selected from the hold-out test set that were independently reviewed by 5 radiologists together with the blood test results. The independent results from radiologists showed an accuracy ranging from 43.7% to 58.7%. This value rose to 59.3% (178/300) when the consensus interpretation of all 5 radiologists based on the majority vote was considered. Of note, the overall accuracy achieved by MultiCOVID was 69.6% (209/300) that was significantly higher than consensus interpretation (P < .001). In addition, for COVID-19 prediction individually, MultiCOVID showed similar sensitivity to the radiologists’ consensus but with a much higher specificity, leading to significantly better performance when discerning between COVID-19 vs Control and COVID-19 vs HF patients (P < .05 for both comparisons; **Figure 5**).

**Figure 5.**
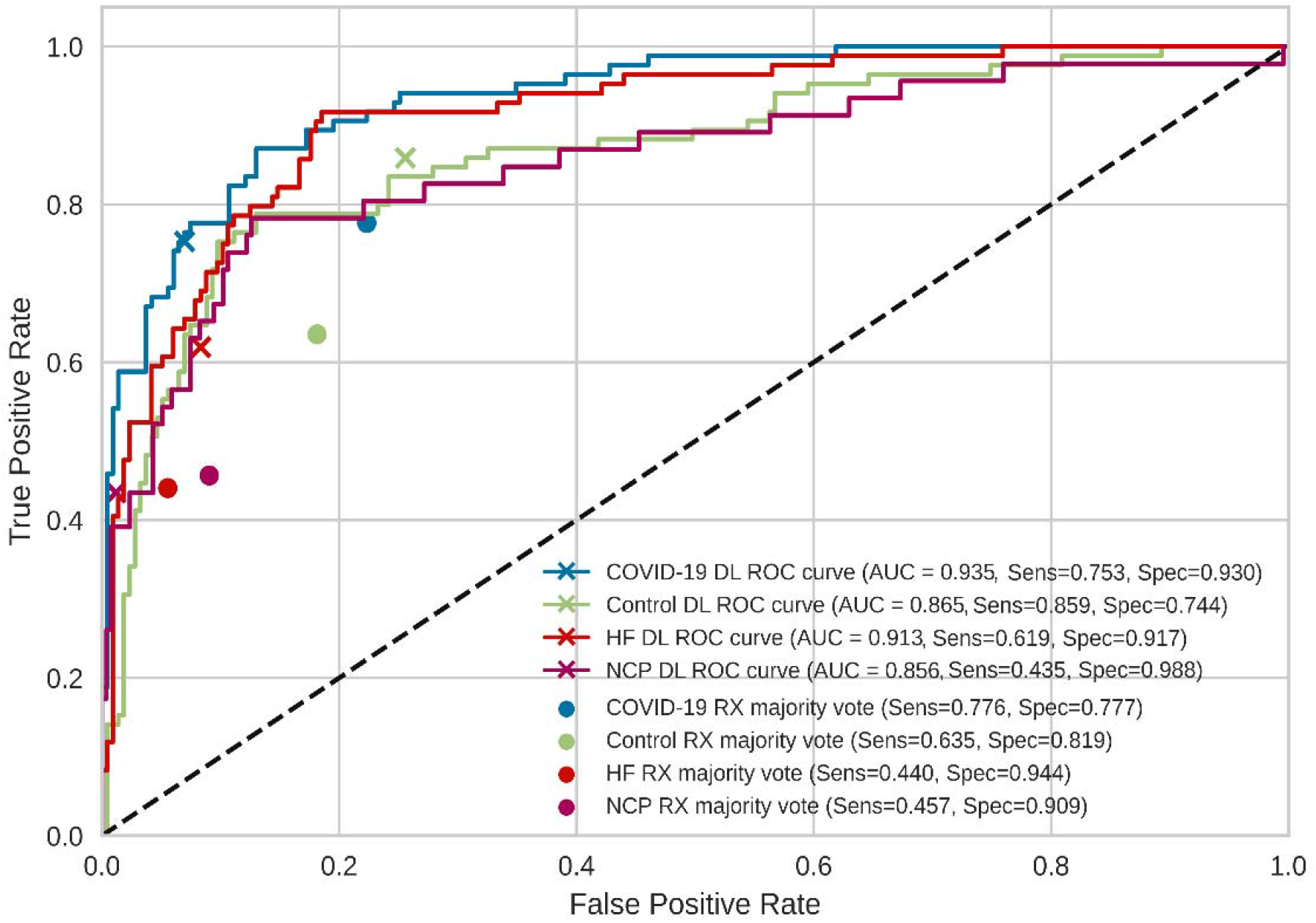
Comparison of the performance of MultiCOVID model with consensus expert radiologist interpretations on random sample of 300 images from the test set. The receiver operating characteristic (ROC) curves for each category (COVID-19 – blue; Control – green; Heart Failure (HF) – red and Non-COVID Pneumonia (NCP) – magenta) are shown for MultiCOVID (DL) and for the consensus interpretation of radiologists (majority vote). Sensitivity (Sens) and specificity (Spec) are also plotted for each category assessed. DL: deep learning

## 4. Discussion

Diagnosis of COVID-19 is an evolving challenge. During the beginning of the pandemic and the successive peaks with high prevalence rates, a prompt and effective diagnosis was critical for proper patient isolation and evaluation. However, since the prevalence of the COVID-19 cases oscillated, showing fewer cases between waves, and more non-covid cases, it was important to differentiate patients with other diseases than COVID-19 presenting similar visual characteristics in the CXR.

During patient assessment in the emergency room, clinicians take into account different inputs for a proper diagnosis. First, the anamnesis, symptoms, vitals and physical findings guide the physician to an initial assumption. Based in this information, additional tests are requested (CXR, blood test, ECG and SARS-CoV-2 detection). The integration of these results allows the team to diagnose a patient accurately. However, this process is time consuming and sometimes findings are difficult to interpret leading to misdiagnosis.

To improve this diagnostic process, we have developed and trained a multimodal deep learning algorithm based in a multiple input approach combining CXR images together with blood sample data to identify COVID-19 diagnosis with high sensitivity. This way we were able to manage the increased complexity of the dataset. These data from multiple sources are somehow correlated and complementary to each other and could reflect patterns that are not present in single models alone (Ngiam et al., 2011).

Hence, MultiCOVID is fed by two of the most common and fast clinical tests requested in the emergency room (CXR and Blood test) and can predict the presence of three different diseases (COVID-19, heart failure and non-COVID19 pneumonia) with similar CXR characteristics.

Analysis of single models shows the importance of model interpretation. While CXR-only models could identify patterns outside the lung parenchyma that could diminish its generalization capacity (Wang et al., 2019), Blood-only models could point to interesting population of cells that are differently represented in COVID-19 patients, leveraging its prediction capacity. In this sense, the immune compartment plays an important role in the COVID-19 response and it has been already published that COVID-19 patients present fewer overall leukocytes counts and, more concretely, eosinophil counts (Rahman et al., 2021; Tan et al., 2021). Furthermore, oxygen transport seems to be somehow affected, modulating the red cell population. In this regard, in our work we found significant differences in the erythrocyte count and the hemoglobin concentration. Although most of the studies correlate the reduction of this values to severe COVID-19 patients (Lippi and Mattiuzzi, 2020), this is the first dataset to compare them in these four different categories at the time of diagnosis.

Moreover, although a huge amount of literature about COVID-19 diagnosis has been published using only blood tests (Bayat et al., 2021; Chen et al., 2021; Kukar et al., 2021; Soltan et al., 2021) or CXR (Hwang et al., 2019; Wang et al., 2020; Wehbe et al., 2021; Zhang et al., 2021), to our knowledge this is the first study that combines both parameters and compares its prediction capacity. It is clear that merging both sources of data leads to a better prediction performance when compared with the two single models alone and that this difference is more pronounced where the number of cases is scarce. It is important to stress that this combination of data sources addresses the variable prevalence of COVID-19 cases during the pandemic, which is an issue that could not be solved in previous studies (Wehbe et al., 2021; Zhang et al., 2021).

Our study had two main limitations. First, the algorithm was evaluated on a single center; thus, there was likely some degree of bias. Additionally, the sample collection was performed in different time periods for each group of patients, which could present some kind of differences in the CXR image acquisition although this was partially solved using the lung segmentation model.

## 5. Conclusions

We have developed a multimodal deep learning algorithm, MultiCOVID, that discriminates among COVID-19, heart failure, non-covid pneumonia and healthy patients using both CXR and blood test with a significantly better performance than experienced thoracic radiologists.

Our approach and results suggest an innovative scenario where COVID-19 prediction could be identified from other similar diseases and facilitate triage within the emergency room in a COVID-19 low prevalence situation.

## Data Availability

All data produced in the present study are available upon reasonable request to the authors

https://github.com/Tato14/MultiCOVID

## Abbreviations

DL: deep learning
CXR: chest X-rays
AUC: area under the receiver operating characteristic curve
COVID-19: coronavirus disease 2019
RT-PCR: reverse-transcription polymerase chain reaction
SARS-CoV-2: severe acute respiratory syndrome coronavirus 2
HF: heart failure
NCP: non-covid pneumonia

## Author contributions

**Max Hardy-Werbin:** Data curation, Validation, Formal analysis, Investigation, Project administration, Supervision, Roles/Writing - original draft, Writing - review & editing; José **Maria Maiques:** Data curation, Formal analysis, Investigation, Validation, Roles/Writing - original draft, Writing - review & editing; **Marcos Busto:** Data curation, Formal analysis, Investigation Validation, Roles/Writing - original draft, Writing - review & editing; **Isabel Cirera**: Data curation, Validation, Roles/Writing - original draft, Writing - review & editing; **Alfons Aguirre**: Data curation, Validation, Roles/Writing - original draft, Writing - review & editing; **Nieves Garcia-Gisbert**: Investigation, Visualization, Project administration, Roles/Writing - original draft, Writing - review & editin; **Flavio Zuccarino**: Validation, Roles/Writing - original draft, Writing - review & editing; **Santiago Carbullanca**: Validation, Roles/Writing - original draft, Writing - review & editing, **Luis Alexander Del Carpio**: Validation, Roles/Writing - original draft, Writing - review & editing, **Didac Ramal**: Validation, Roles/Writing - original draft, Writing - review & editing; **Ángel Gayete**: Validation, Roles/Writing - original draft, Writing - review & editin; **Jordi Martínez-Roldan**: Project administration, Supervision, Roles/Writing - original draft, Writing - review & editing; **Albert Marquez-Colome**: Data curation, Project administration, Supervision, Roles/Writing - original draft, Writing - review & editing; **Beatriz Bellosillo**: Data curation, Formal analysis, Investigation, Project administration, Supervision, Roles/Writing - original draft, Writing - review & editin; **Joan Gibert**: Data curation, Formal analysis; Investigation, Visualization, Project administration, Supervision, Roles/Writing - original draft, Writing - review & editing.

