## Supplemental Table for "MultiCOVID: a multi modal Deep Learning approach for COVID-19 diagnosis"

**Supplementary Table.** Performance of different models on entries from hold-out test datasets. Metrics are defined as: Precision = True Positive / (True Positive + False Positive); Sensitivity = True Positives / (True Positives + False Negatives); F1 score = mean (Precision, Sensitivity); Accuracy = (True Positives + True Negatives) / (Actual Positives + Actual Negatives). The higher values for each dataset are marked in bold.

| Label | Model | Precision | | Sensitivity | | F1 Score | | Accuracy | | AUC | |
| --- | --- | --- | --- | --- | --- | --- | --- | --- | --- | --- | --- |
|  |  | Mean | SD | Mean | SD | Mean | SD | Mean | SD | Mean | SD |
| COVID-19 | CXR-Only | 65.2% | 6.8% | 64.6% | 5.4% | 64.8% | 4.8% | 79.1% | 0.6% | 91.7% | 1.0% |
| COVID-19 | Blood-Only | 70.4% | 0.5% | 57.1% | 1.3% | 63.0% | 1.0% | 74.3% | 0.2% | **94.9%** | **0.1%** |
| COVID-19 | XGB Blood-Only | 76.8% | 1.2% | 66.8% | 1.9% | 71.5% | 1.2% | 76.9% | 0.5% | 94.6% | 0.8% |
| COVID-19 | Join | 68.4% | 4.0% | 74.2% | 6.1% | 70.9% | 1.6% | 80.4% | 1.3% | 92.0% | 1.6% |
| COVID-19 | MultiCOVID | **76.9%** | **1.0%** | **80.4%** | **1.5%** | **78.6%** | **0.9%** | **84.0%** | **0.5%** | 94.3% | 0.3% |
| Control | CXR-Only | 85.9% | 1.8% | 90.2% | 1.8% | 87.9% | 0.1% | 79.1% | 0.6% | 90.2% | 1.0% |
| Control | Blood-Only | 77.5% | 0.2% | 91.8% | 0.1% | 84.1% | 0.1% | 74.3% | 0.2% | 81.1% | 0.1% |
| Control | XGB Blood-Only | 80.4% | 0.5% | **92.2%** | **0.4%** | 85.9% | 0.2% | 76.9% | 0.5% | 84.2% | 0.7% |
| Control | Join | 89.1% | 1.6% | 88.6% | 2.8% | 88.8% | 1.1% | 80.4% | 1.3% | 88.7% | 2.5% |
| Control | MultiCOVID | **89.9%** | **0.1%** | 91.7% | 0.6% | **90.8%** | **0.3%** | **84.0%** | **0.5%** | **90.6%** | **0.5%** |
| HF | CXR-Only | 57.1% | 4.1% | 50.3% | 11.7% | 52.9% | 7.2% | 79.1% | 0.6% | 91.4% | 1.4% |
| HF | Blood-Only | 36.7% | 1.0% | 14.9% | 0.9% | 21.3% | 1,0% | 74.3% | 0.2% | 77.0% | 0.1% |
| HF | XGB Blood-Only | 45.1% | 3.0% | 27.2% | 3.0% | 33.9% | 2.9% | 76.9% | 0.5% | 82.4% | 0.9% |
| HF | Join | 55.6% | 2.6% | 56.8% | 8.0% | 55.9% | 4.1% | 80.4% | 1.3% | 88.2% | 2.2% |
| HF | MultiCOVID | **61.0%** | **2.1%** | **59.6%** | **2.2%** | **60.3%** | **2.0%** | **84.0%** | **0.5%** | **92.4%** | **0.3%** |
| NCP | CXR-Only | 55.8% | 6.1% | 35.0% | 5.8% | 42.8% | 5.5% | 79.1% | 0.6% | 86.0% | 2.2% |
| NCP | Blood-Only | 49.5% | 1.2% | 18.8% | 0.0% | 27.2% | 0.2% | 74.3% | 0.2% | 78.6% | 0.1% |
| NCP | XGB Blood-Only | 28.1% | 13.1% | 5.2% | 1.9% | 8.7% | 3.3% | 76.9% | 0.5% | 72.1% | 1.7% |
| NCP | Join | 52.2% | 7.0% | 41.7% | 14.1% | 45.1% | 8.2% | 80.4% | 1.3% | 83.5% | 7.1% |
| NCP | MultiCOVID | **68.2%** | **4.2%** | **48.3%** | **4,0%** | **56.5%** | **4.1%** | **84.0%** | **0.5%** | **89.2%** | **0.4%** |

Abbreviations: COVID-19: coronavirus disease 2019, HF: heart failure, NCP: non-covid pneumonia, CXR: Chest X-Ray, AUC: area under the curve.
